# An incomplete Circle of Willis is not a risk factor for white matter hyperintensities: The Tromsø Study

**DOI:** 10.1101/2020.09.25.20183756

**Authors:** Lars B. Hindenes, Asta K. Håberg, Ellisiv B. Mathiesen, Torgil R. Vangberg

**Affiliations:** Department of Clinical Medicine, Faculty of Health Sciences, UiT The Arctic University of Norway, Postboks 6050 Langnes 9037 Tromsø, Norway; PET Centre, University Hospital of North Norway, 9038 Tromsø, Norway; Department of Radiology and Nuclear Medicine, St. Olav University Hospital, Postboks 3250 Torgarden 7030 Trondheim, Norway; Department of Neuromedicine and Movement Science, Norwegian University of Science and Technology (NTNU), NTNU 7491 Trondheim, Norway; Department of Neurology, University Hospital of North Norway, 9038 Tromsø, Norway

**Keywords:** Circulation, Leukoaraiosis, Health survey, Epidemiology

## Abstract

**Objective:** The Circle of Willis (CoW) is often underdeveloped or incomplete, leading to suboptimal blood supply to the brain. As hypoperfusion is thought to play a role in the aetiology of white matter hyperintensities (WMH), the objective of this study was to assess whether incomplete CoW variants were associated with increased WMH volumes compared to the complete CoW.

**Methods:** In a cross-sectional population sample of 1864 people (age 40 – 84 years, 46.4% men), we used an automated method to segment WMH using T1-weighted and T2-weighted fluid-attenuated inversion recovery image obtained at 3T. CoW variants were classified from time-of-flight scans, also at 3T. WMH risk factors, including age, sex, smoking and blood pressure, were obtained from questionnaires and clinical examinations. We used linear regression to examine whether people with incomplete CoW variants had greater volumes of deep WMH (DWMH) and periventricular WMH (PWMH) compared to people with the complete CoW, correcting for WMH risk factors.

**Results:** Participants with incomplete CoW variants did not have significantly higher DWMH or PWMH volumes than those with complete CoW when accounting for risk factors. Age, pack-years smoking, and systolic blood pressure were risk factors for increased DWMH and PWMH volume. Diabetes was a unique risk factor for increased PWMH volume.

**Conclusion:** Incomplete CoW variants do not appear to be risk factors for WMH in the general population.

## 1. Introduction

White matter hyperintensities (WMH) are seen as a marker of cerebral small vessel disease (Wardlaw et al., 2015). They are common in older adults, and their prevalence rapidly increases with age (de Leeuw, 2001). WMH have been associated with cognitive decline and dementia (Debette and Markus, 2010), and impaired motor function (Fleischman et al., 2015). Risk factors of cerebrovascular disease have also been associated with WMH, including hypertension (Dufouil et al., 2001; van Dijk et al., 2008, 2004), type 1 and 2 diabetes (Habes et al., 2016; King et al., 2014), body mass index (BMI) (King et al., 2014), and smoking (Gons et al., 2011; Habes et al., 2016; van Dijk et al., 2008). Although the aetiology of WMH is not fully understood (Wardlaw et al., 2015), hypoperfusion may play a role as histology shows signs of hypoxia in WMH (Fernando et al., 2006) and WMH commonly occur in watershed regions in cerebral white matter.

The Circle of Willis (CoW), located at the base of the brain, is an anastomosis of the carotid arteries and the basilar artery. Its circular structure enables redistribution of the cerebral blood flow in case of reduced upstream flow, subsequently providing redundancy to the brain’s blood supply. However, it is very common that at least one of the segments in the CoW are hypoplastic or missing. A recent dissection study found that only 7% of adults have a complete CoW (Wijesinghe et al., 2020), suggesting that the CoW does not inherently have a critical influence on human survival. A non-patent CoW may, however, cause suboptimal blood supply to some brain regions due to diminished collateral capacity, particularly in combination with atherosclerosis or other arterial occlusions.

Indeed, previous studies have found an association between incomplete CoW variants and increased WMH burden. In a convenience sample of 163 patients, it was found that patients with incomplete variants had a significantly higher Fazekas score compared to those with a complete CoW (Ryan et al., 2015). In patients with carotid artery stenosis, incomplete CoW variants were associated with greater WMH volume and number of WMH lesions (Saba et al., 2017, 2015), increased WMH rating (Chuang et al., 2011), and increased deep WMH (DWMH) and periventricular WMH (PWMH) ratings (Ye et al., 2019). However, the results are not unison, as some studies fail to find a link between more WMH and decreased completeness of the CoW, both in patients with carotid stenosis (Li et al., 2015; van der Grond et al., 2004) and in the general population (Del Brutto and Lama, 2015). Despite diverging results from prior studies, there is evidence suggesting that an incomplete CoW may be a risk factor for WMH possibly via impaired autoregulation of cerebral blood flow (Guo et al., 2018) which has been shown to be associated with WMH (Purkayastha et al., 2014). Furthermore, PWMH is more strongly associated with cerebral blood flow (ten Dam et al., 2007), blood pressure (Griffanti et al., 2018) and vasculature in general (Armstrong et al., 2020) than DWMH. Therefore, PWMH may be more sensitive to variations in the CoW than DWMH, and this may in part explain possible diverging findings relating to WMH and anatomical variants of the CoW. Since WMH and also incomplete CoW variants (El-Barhoun et al., 2009; Hindenes et al., 2020; Zaninovich et al., 2017) are more prevalent among older people, it is of interest to examine whether incomplete CoW variants pose a risk of WMH in the general population.

In this study, we assessed whether incomplete CoW variants might be risk factors for increased WMH burden in a large population-based sample of middle-aged and older people. We used linear regression models to compare DWMH and PWMH volumes of each incomplete CoW variant to the complete variant while correcting for cerebrovascular risk factors.

## 2. Methods

The study was approved by the Regional Committee of Medical and Health Research Ethics Northern Norway (2014/1665/REK-Nord) and carried out in accordance with relevant guidelines and regulations at UiT The Arctic University of Norway. All participants gave written informed consent before participating in the study.

### 2.1. Study population

Eligible for the study were 1864 participants (40 – 84 years) of the seventh wave of the population-based Tromsø Study (Njølstad et al., 2016) who underwent time-of-flight (TOF) MRI scanning of the brain and had complete imaging data (Hindenes et al., 2020). We excluded participants with image artefacts or brain pathology that could lead to unreliable WMH volume estimates, and participants with rare CoW variants (less than ten observations) as parameter estimates would be unreliable.

### 2.2. MRI protocol

Participants were scanned at the University Hospital North Norway with the same 3T Siemens Skyra MR scanner (Siemens Healthcare, Erlangen, Germany). We used a 64-channel head coil in the majority of examinations, but in 38 examinations a slightly larger 20-channel head coil had to be used to accommodate the participants’ head. The MRI protocol consisted of T1-weighted (T1w), T2-weighted fluid-attenuated inversion recovery (FLAIR), TOF angiography and susceptibility-weighted sequences.

T1w images were acquired with a 3D magnetisation prepared rapid acquisition gradient-echo (MPRAGE) sequence (flip angle=9°, TR/TE/TI=2300 ms/4.21 ms/996 ms). The T2-weighted FLAIR images were acquired with a 3D turbo spin-echo sequence with variable flip angle (TR/TE/TI = 5000 ms/388 ms/1800 ms, partial Fourier = 7/8). The T1w and FLAIR scans were acquired sagittally with 1 mm isotropic resolution, Generalized Autocalibrating Partially Parallel Acquisition (GRAPPA) parallel imaging acceleration factor 2, FOV = 256 mm, 176 slices, 1 mm slice thickness, and 256 x 256 image matrix. The T1w and FLAIR were used to measure the WMH volume. TOF angiography images were acquired with a 3D transversal fast low angle shot (FLASH) sequence with flow compensation (TR/TE = 21/3.43 ms, GRAPPA parallel imaging acceleration factor 3, FOV 200 x 181 mm, slice thickness 0.5 mm, 7 slabs with 40 slices each). Reconstructed image resolution was 0.3 x 0.3 x 0.5 mm.

### 2.3. Defining Circle of Willis segments and mirrored variants

The CoW consists of the left and right proximal anterior cerebral artery (A), the anterior communicating artery (Ac), the left and right posterior communicating artery (Pc), and the left and right proximal posterior cerebral artery (P). In figures and tables, we refer to the complete CoW variant as “O”, while the incomplete variants are denoted by their missing segments. We did not differentiate between left-right mirrored CoW variants in order to reduce the number of tested variants. Including the complete variant, we had in total 17 unique CoW variants (Fig. 1). The classification of anatomical variants in the CoW were done in a previous study, where segments not visible or with a diameter less than 1 mm were classified as missing/hypoplastic (Hindenes et al., 2020).

**Fig. 1.**
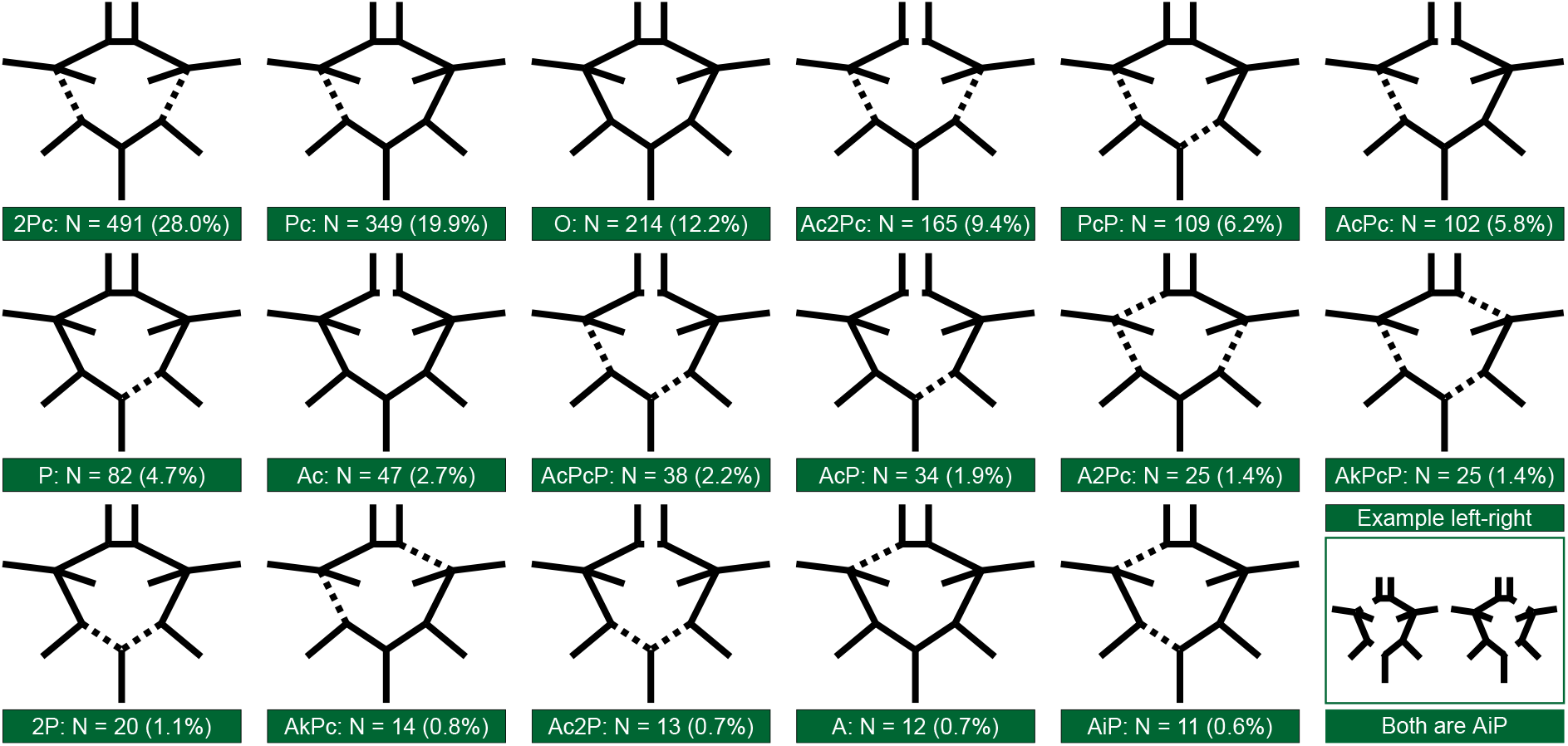
The 17 Circle of Willis variants used in the analyses. “N” and “%” denotes count and percentage of each variant. If asymmetric, each schematic Circle of Willis and label contain both left and right variants as shown by the example in the lower right corner. “i” = ipsilateral and “k” = contralateral relationship between two subsequent segments. Prefix “2” denotes that both left and right segments are missing. “A” = proximal anterior cerebral artery, “Ac” = anterior communicating artery, “Pc” = posterior communicating artery, “P” = proximal posterior cerebral artery, and “O” = complete Circle of Willis.

### 2.4. WMH segmentation

We segmented WMH using a fully convolutional neural network algorithm (UNET) (Li et al., 2018) using the T1w and FLAIR images. This algorithm was the best performer in the WMH Segmentation Challenge at MICCAI 2017 (Kuijf et al., 2019) and was trained on images fromfive different scanners. The algorithm should therefore be more robust for use on images from a different scanner.

The T1w and FLAIR images were first co-registered, and then rigid body aligned to the MNI template using ANTs toolkit (http://stnava.github.io/ANTs/, v.2.3.1). The latter was done to ensure consistent orientation of the images, which were found to improve the reliability of the UNET algorithm. We ran the UNET algorithm following Li’s and colleagues’ recommendations and with default settings, except that we did not use bias-field correction, as an initial validation showed that this degraded the segmentation. After UNET, the segmentation was corrected for misclassifications in grey matter by applying a white matter mask derived from the FreeSurfer (v6.0) subcortical segmentation of the T1w images (Fischl et al., 2004, 2002).

We validated the WMH segmentation against 30 manually segmented images with a wide range of WMH volumes, from almost no WMH to severe volumes. The automatic segmentation showed similar performance in terms of the similarity coefficient Dice (mean Dice = 0.519 [SD = 0.234]) compared to many other available WMH segmentation methods (Griffanti et al., 2016; Jiang et al., 2018; Kamnitsas et al., 2017; Rachmadi et al., 2018; Schmidt et al., 2012; Valverde et al., 2017). Note that the Dice score becomes more sensitive to errors at small volumes, thereby decreasing the mean Dice score for the validation dataset, but for participants with WMH volumes > 2.5 ml (2500 voxels) the Dice scores were almost always greater than the mean Dice score, averaging close to 0.7, see Fig. 2.

**Fig. 2.**
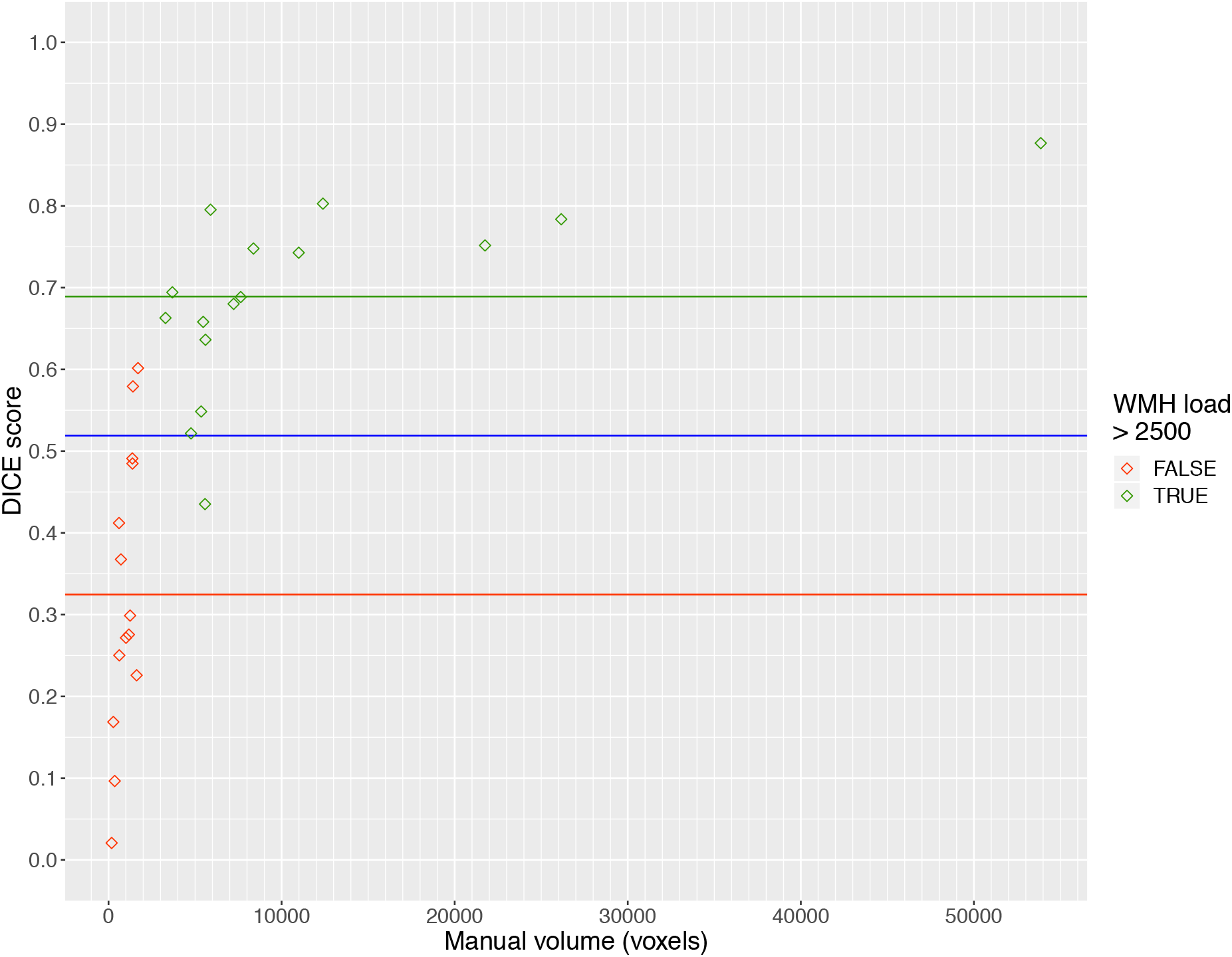
Validation of the WMH segmentation algorithm: Dice score and relative absolute volume difference between ground truth WMH and segmented WMH. The blue line shows the overall mean Dice score, while the green and red line show the mean Dice score for segmentations with respective ground truth WMH load over and under 2500 voxels.

### 2.5. Partitioning WMH into deep and periventricular segments

We partitioned the WMH masks into PWMH and DWMH using a 10 mm distance from the ventricles. We chose 10 mm as a threshold based on a recent study (Griffanti et al., 2018) which showed that this threshold gives the most comparable results to previous studies and also the best separation between PWMH and DWMH in terms of the cardiovascular risk factors. Masks separating PWMH and DWMH were created by spherically dilating ventricle masks from the FreeSurfer segmentation by 10 mm.

### 2.6. Demographic and WMH risk variables

We included ten adjustment variables in our models that have been associated with WMH: age (Nyquist et al., 2015; van Dijk et al., 2008), sex (Nyquist et al., 2015; Rostrup et al., 2012), BMI (King et al., 2014), education (Habes et al., 2016), smoking (Gons et al., 2011; Habes et al., 2016; van Dijk et al., 2008) as pack-years, diabetes (Habes et al., 2016; King et al., 2014), blood glucose levels (Cherbuin et al., 2015), high-sensitive C-reactive protein (CRP) levels (Satizabal et al., 2012), systolic blood pressure (SBP) (Dufouil et al., 2001; van Dijk et al., 2008, 2004), and total cholesterol to high-density lipoprotein (HDL) ratio (Dickie et al., 2016). The independent and dependent variables are further characterised in Table 1. Missing values in continuous variables were imputed by mean or median depending on their distribution, and missing values in categorical variables were imputed by rounded median.

### 2.7. Statistical analysis

All statistical analyses were done with R (v3.5.2). In the linear regression analyses, we used (1) DWMH volume, and (2) PWMH volume as dependent variables. Both volume estimates were log-transformed to better satisfy normality and homoscedasticity assumptions, and we added 1 voxel (1 cubic mm) to all volumes before the log transformation to avoid log of zero. For each dependent variable, we fitted a model including age, sex, CoW variants, and risk factors; i.e. each model has ten risk factors, and 17 factors for each of the incomplete CoW variants including the complete CoW used as a common reference.

Post-hoc multiple comparisons of means with Dunnett contrasts were used to test whether the incomplete CoW variants had higher WMH volumes compared to the complete CoW variant (i.e. a one-sided test). This was done separately for each model. Each post-hoc comparison reports step-down Dunnett adjusted p-values relative to the conventional significance level for one-sided comparisons. The R package “multcomp” (Hothorn et al., 2008) was used to calculate the post-hoc comparisons with adjusted p-values based on the linear regression models. The corresponding confidence intervals constructed to calculate p-values were stochastic, making exact replication without a seed unlikely. To avoid an unwarranted increase in the probability of committing type 1 errors, at the cost of slightly increased risk of type 2 error, we corrected only for multiple tests within linear regression models as described, but not between models. We regarded the conventional p < 0.05 as significant.

## 3. Results

### 3.1. Study participants

Of the 1864 participants, 76 were excluded due to atrophy or image artefacts that might have led to inaccurate WMH volume estimates, 26 were excluded due to having a rare CoW variant, and 11 were excluded due to failures in the segmentation algorithm (see Fig. 3). Mean age for all participants was 63.5 years (SD = 10.6). Men were on average 64.3 years (SD = 10.5), and women 62.8 years (SD = 10.7). Further sample characteristics are shown in Table 1.

**Fig. 3.**
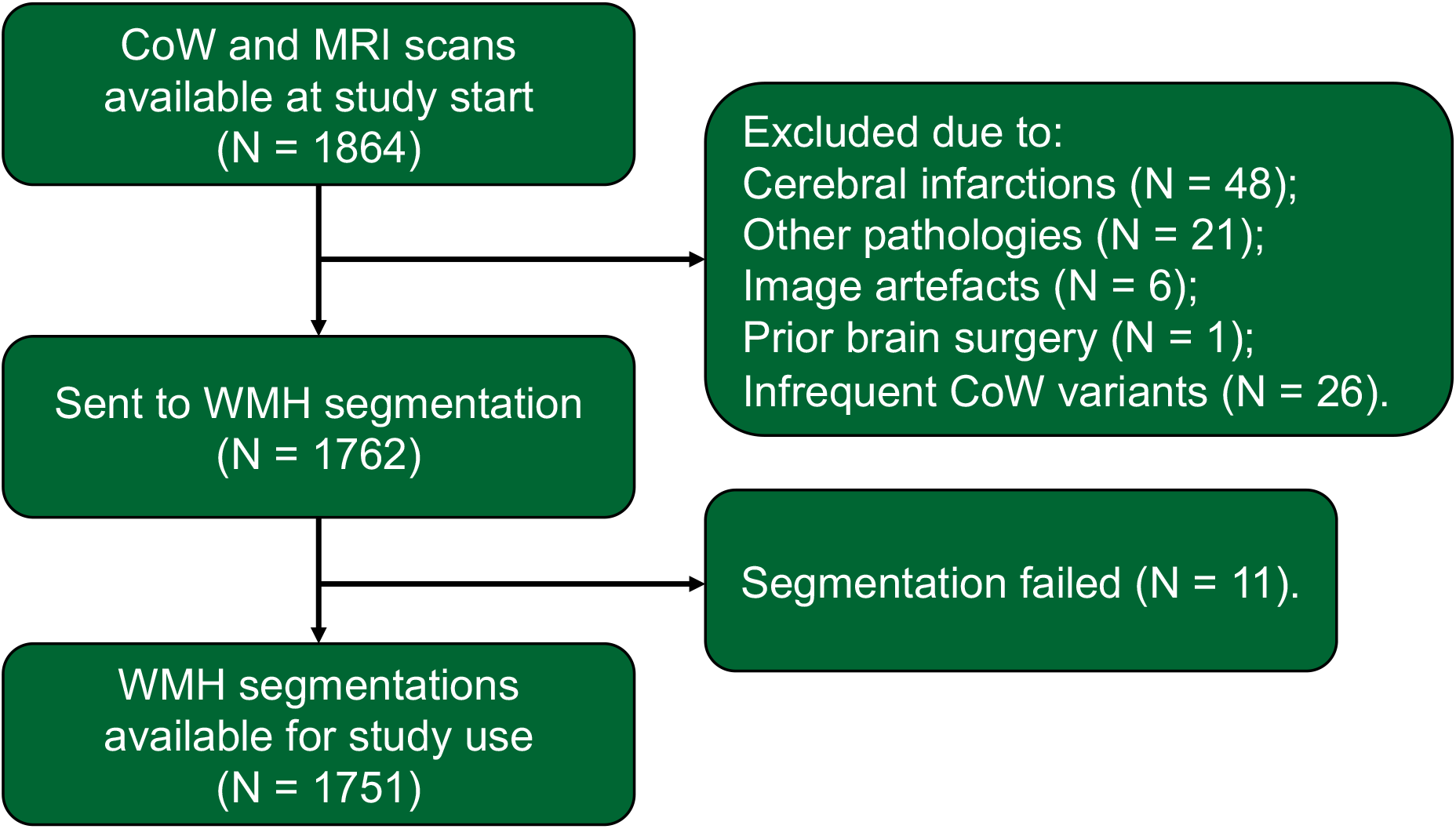
Flow diagram of the sample selection process. “CoW” = Circle of Willis, “WMH” = White matter hyperintensities.

**Table 1.** Characteristics of the study participants.

| Variable | Unit or Factor | Measurement | Missing* | Min | Max |
| --- | --- | --- | --- | --- | --- |
| **Age** | Year | 63.51 (10.63) | 0 | 40.00 | 84.00 |
| **Sex** | Female | 938 (53.6) | 0 |  |  |
|  | Male | 813 (46.4) |  |  |  |
| **Education** | Up to 10 years | 509 (29.7) | 37 |  |  |
|  | High school: 3+ years | 471 (27.5) |  |  |  |
|  | Up to 4 years at university | 340 (19.8) |  |  |  |
|  | More than 4 years at university | 394 (23.0) |  |  |  |
| **SBP** | mmHg | 133.96 (20.79) | 6 | 78.00 | 220.00 |
| **Hypertension<sup>#</sup>** | No | 1070 (61.3) | 6 |  |  |
|  | Yes | 675 (38.7) |  |  |  |
| **Cholesterol to HDL ratio** |  | 3.67 (1.35) | 7 | 1.28 | 23.00 |
| **CRP, median [IQR]** | mg/L | 1.02<br>[0.59, 2.08] | 7 | 0.14 | 58.98 |
| **Smoke daily** | Yes, now | 222 (12.8) | 18 |  |  |
|  | Yes, previously | 839 (48.4) |  |  |  |
|  | Never | 672 (38.8) |  |  |  |
| **Pack years** |  | 9.39 (12.87) | 71 | 0.00 | 90.00 |
| **BMI** |  | 27.15 (4.20) | 1 | 13.90 | 45.90 |
| **Diabetes** | No | 1587 (93.6) | 55 |  |  |
|  | Yes | 109 (6.4) |  |  |  |
| **Glucose** | mmol/L | 5.74 (1.65) | 3 | 2.70 | 19.20 |
| **Total WMH, median [95% CI]** | mm<sup>3</sup> | 2113 [505, 31584] | 0 | 0 | 110447 |
| **DWMH, median [95% CI]** | mm<sup>3</sup> | 699 [221, 9734] | 0 | 0 | 41634 |
| **PWMH, median [95% CI]** | mm<sup>3</sup> | 1332 [166, 21725] | 0 | 0 | 74558 |

Unless specified, continuous variables are given as mean (standard deviation), and
categorical variables as count (percentage). Abbreviations: WMH = white matter
hyperintensities, DWMH = deep white matter hyperintensities, PWMH = periventricular
white matter hyperintensities, CRP = C-reactive protein, SBP = systolic blood pressure, MMS
= mini mental status, BMI = body mass index, HDL = high density lipoprotein, CI = credible
interval, IQR = interquartile range, and SD = standard deviation. \* Missing values were
imputed before analysis. # Hypertension were defined as systolic blood pressure $\geq 140$
mmHg or diastolic blood pressure $\geq 90$ mmHg.

### 3.2. Deep WMH model with post-hoc multiple comparisons

There was no incomplete CoW variant with significantly higher DWMH volume compared to the complete CoW variant in the post-hoc multiple comparisons (Table 2). The PcP variants were the only CoW with an adjusted p-value under 50% (t = 2.457, adjusted p = 0.093) and its unadjusted equivalent two-sided p-level was significant at 0.05. Altogether this suggests that the PcP variants trended to have more DWMH than the complete variant. The R^2^ for the regression model was 0.167, indicating that this model did not explain DWMH well.

**Table 2.**
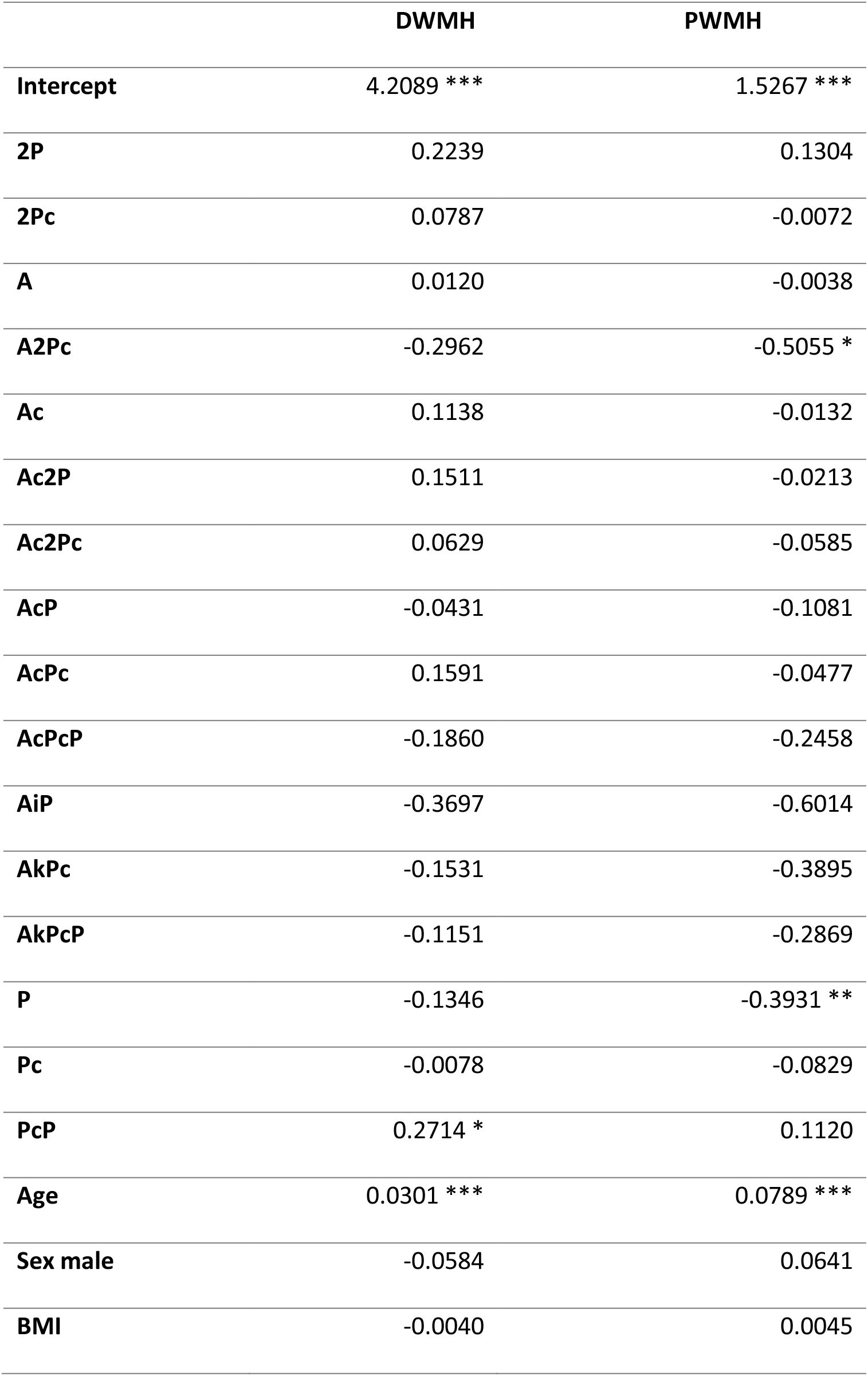
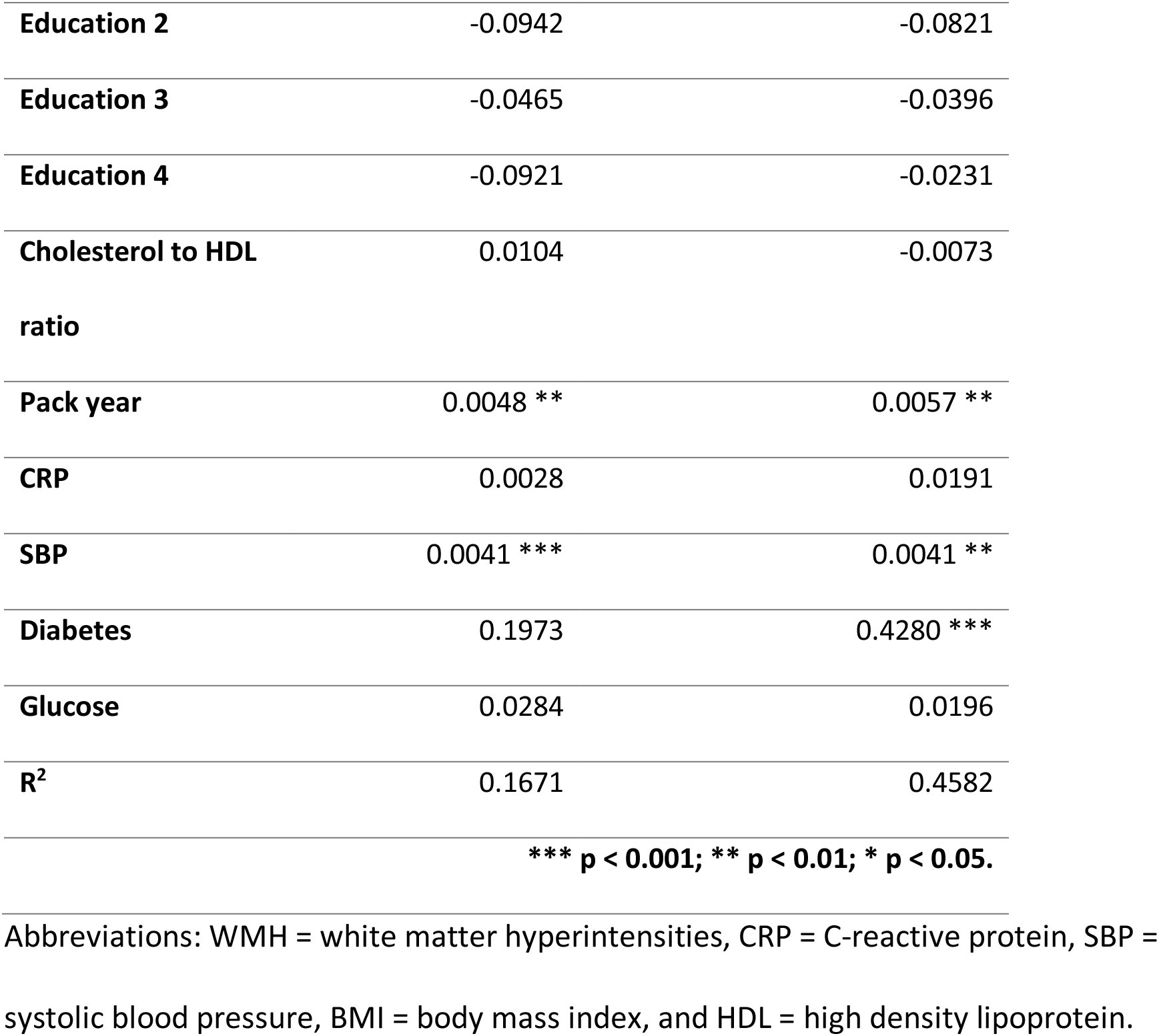
Regression model coefficients for DWMH and PWMH. p-levels denoted by asterisk in the table are from unadjusted two-sided tests in the models included for gauging risk of type 1 error, and not results of the one-sided post-hoc analysis.

|  | **DWMH** | **PWMH** |
| --- | --- | --- |
| **Intercept** | 4.2089 *** | 1.5267 *** |
| **2P** | 0.2239 | 0.1304 |
| **2Pc** | 0.0787 | -0.0072 |
| **A** | 0.0120 | -0.0038 |
| **A2Pc** | -0.2962 | -0.5055 * |
| **Ac** | 0.1138 | -0.0132 |
| **Ac2P** | 0.1511 | -0.0213 |
| **Ac2Pc** | 0.0629 | -0.0585 |
| **AcP** | -0.0431 | -0.1081 |
| **AcPc** | 0.1591 | -0.0477 |
| **AcPcP** | -0.1860 | -0.2458 |
| **AiP** | -0.3697 | -0.6014 |
| **AkPc** | -0.1531 | -0.3895 |
| **AkPcP** | -0.1151 | -0.2869 |
| **P** | -0.1346 | -0.3931 ** |
| **Pc** | -0.0078 | -0.0829 |
| **PcP** | 0.2714 * | 0.1120 |
| **Age** | 0.0301 *** | 0.0789 *** |
| **Sex male** | -0.0584 | 0.0641 |
| **BMI** | -0.0040 | 0.0045 |
| **Education 2** | -0.0942 | -0.0821 |
| **Education 3** | -0.0465 | -0.0396 |
| **Education 4** | -0.0921 | -0.0231 |
| **Cholesterol to HDL ratio** | 0.0104 | -0.0073 |
| **Pack year** | 0.0048 ** | 0.0057 ** |
| **CRP** | 0.0028 | 0.0191 |
| **SBP** | 0.0041 *** | 0.0041 ** |
| **Diabetes** | 0.1973 | 0.4280 *** |
| **Glucose** | 0.0284 | 0.0196 |
| **R<sup>2</sup>** | 0.1671 | 0.4582 |
| *** p < 0.001; ** p < 0.01; * p < 0.05. |  |  |

Abbreviations: WMH = white matter hyperintensities, CRP = C-reactive protein, SBP = systolic blood pressure, BMI = body mass index, and HDL = high density lipoprotein.

### 3.3. Periventricular WMH model with post-hoc multiple comparisons

For the PWMH model (Table 2), no incomplete CoW variants had significantly higher PWMH volume compared to the complete CoW variant in the post-hoc multiple comparisons analysis. All adjusted p-values were between 0.85 – 1. In terms of the two-sided p-levels, the P and A2Pc variants had negative coefficients significant at 0.01 and 0.05, respectively, explaining why the one-sided post-hoc analysis was non-significant. The R^2^ for the regression model was 0.458, and also considerably higher than for the DWMH model. This indicates that the same set of vascular risk factors and incomplete CoW variants in the regression models explained PWMH volume better than DWMH volume.

## 4. Discussion

In this population-based study, we examined whether participants with incomplete CoW variants had increased WMH volumes compared to those with the complete CoW. The main findings were that neither PWMH or DWMH volumes were significantly increased in participants with incomplete CoW variants relative to participants with a complete CoW when correcting for age, gender and WMH risk factors. To our knowledge, only one previous study has examined whether there is an association between the completeness of the CoW and WMH in the general population (Del Brutto and Lama, 2015). This study examined older participants from a native South American population, that have a high prevalence of SVD, but failed to find any association between an incomplete CoW and increased WMH (Del Brutto and Lama, 2015) and markers of SVD in general (Del Brutto et al., 2015). Our results, together with these findings suggest that anatomical variations in the CoW are not substantial risk factors for increased DWMH and PWMH volumes in the general population aged 40 years or older.

Our results differ from previous studies that have found an association between incomplete CoW variants and increased WMH load. Most of these studies are on patients with carotid artery stenosis (Chuang et al., 2011; Saba et al., 2017, 2015; Ye et al., 2019). For this patient group, there is a clear mechanism connecting the CoW anatomy and WMH, i.e., the stenosis will render the collateral capacity of the CoW important for maintaining sufficient blood flow to the brain. The association between the incompleteness of CoW and lower WMH load in atherosclerosis patients together with our negative results on generally healthy participants suggests that collateral flow in the CoW may only be protective against WMH when arteries upstream to the CoW are stenotic or occluded. However, it is difficult to firmly conclude that an incomplete CoW is a risk factor for WMH even in patients with carotid artery stenosis as some studies have failed to find such an association (Li et al., 2015; van der Grond et al., 2004).

The inconsistent results on whether a complete CoW may be protective against WMH may hint of a more complex role of the CoW in the cerebrovascular system than just having the role as a compensatory system in case of arterial occlusion. This has been suggested by previous studies (Pascalau et al., 2018; Vrselja et al., 2014). In particular, Vrselja et al. argued that the CoW also plays a role in blood pressure dissipation, ensuring lower fluctuations in blood pressure in the downstream cerebral arteries of the CoW (Vrselja et al., 2014). It is conceivable that hypoplastic segments with a diameter less than 1 mm may have an important role in dissipating fluctuations in blood pressure, but only a minor role in collateral blood flow. As greater pressure fluctuations in the blood flow are associated with WMH (Purkayastha et al., 2014), the dampening of blood pressure fluctuations in the CoW may be just as crucial as its collateral ability. Unfortunately, we did not distinguish between missing and hypoplastic segments, similarly to most other TOF imaging studies on the CoW (Del Brutto and Lama, 2015; El-Barhoun et al., 2009; Krabbe-Hartkamp et al., 1998; Qiu et al., 2015; Saba et al., 2017), since it is near impossible to distinguish between missing and hypoplastic segments reliably from TOF images.

As expected, some of the WMH risk factors were associated with increased DWMH and PWMH. Interestingly, diabetes was only associated with increased PWMH, lending support to the notion that DWMH and PWMH have differing aetiology (Armstrong et al., 2020; ten Dam et al., 2007). Also, vascular risk factors, age and gender explained 45.8% of the variance in PWMH, while only 16.7% of the variance of DWMH, suggesting that PWMH is more closely associated with cerebrovascular disease than DWMH.

There are some limitations to our study warranting discussion. First, it is conceivable that other measurements of the CoW might offer greater insight into a potential connection between the CoW anatomy and WMH; e.g. considering the diameters of the arterial segments and, as already discussed, distinguishing between hypoplastic and absent segments. Second, we divided the WMH volume into deep and periventricular components which can make our study difficult to compare with other studies. Third, it is possible that partitioning the WMH volume in the individual flow territories would be more sensitivity to variations in the CoW anatomy. Still, we decided against this because flow territories depend on the CoW anatomy (van Laar et al., 2006). Fourth and last, most previous studies on the connection between WMH and CoW have grouped the anatomical variations of the CoW into broad categories (e.g. missing segments in the anterior or posterior circulation), instead of comparing unique variants as we have done. The rationale for this was that it might provide greater sensitivity, but it also complicates the comparison to previous studies.

The strengths of the study are the large cross-sectional population-based sample with reliable information on relevant risk factors. We also used a state of the art automatic WMH segmentation software to avoid WMH rater biases. Use of such automatic software has also increased the replicability of our study and allowed us to use WMH volumes instead of discrete scales, as this is found to offer greater sensitivity in correlations with clinical measures (van Straaten et al., 2006). Our CoW classification also has acceptable intra and inter rater accuracy (Hindenes et al., 2020). Furthermore, due to our large sample we, were able to include 17 unique CoW variants in the models without compromise in model assumptions. We applied the liberal step-down Dunnett correction for multiple tests, compared to Bonferroni, to reduce chance of type 2 errors. We also provided information about regressions’ two-sided p-levels enabling some insight about chance of type 1 errors or whether our post hoc analysis had missed possible negative associations not originally hypothesized as relevant.

## 5. Conclusions

In this large population sample of people aged 40 and older, incomplete CoW variants are not substantial risk factors for increased DWMH and PWMH volumes.

## Data Availability

Researchers can obtain the data used in this study by applying to The Tromsø Study via a standard application procedure (https://en.uit.no/forskning/forskningsgrupper/gruppe?p_document_id=453582).

http://tromsoundersokelsen.uit.no/tromso/

## Funding

This work was supported by two Helse Nord project grants HNF1369-17 and SFP1271-16, and computational resources from NOTUR grant #NN9562K. The funding source had no role in the study design, collection of data, analysis, interpretation of data, and in the decision to submit the article for publication.

## Declaration of Competing Interests

None.

## Acknowledgements

We thank the participants of the Tromsø Study, the administration of the Tromsø Study, the Department of Radiology at the University Hospital North Norway and the MR imaging technologists for their contributions to the study. Special thanks to Liv Hege Johnsen for providing necessary aid in data curation process.

## Notes

### Competing Interest Statement

The authors have declared no competing interest.

